# A Bibliometric Approach to Scientific Production on Familial Hypophosphatemic Rickets in Scopus (2000-2022)

**DOI:** 10.1101/2023.12.26.23300534

**Authors:** Frank Hernández-García, Helena Gil-Peña, Julián Rodríguez Suárez, José Manuel López García, Rocío Fuente Pérez, Patricia Oro Carbajosa, Ibrain Enrique Corrales-Reyes

## Abstract

**Background:** Hypophosphatemic rickets are disabling conditions that negatively impact physical functioning, activities of daily living, mental health, social life, and leisure activities. The most common cause of Hypophosphatemic rickets is genetic factors, such as X-linked hypophosphatemia. The evaluation of the scientific application of familial hypophosphatemic rickets aids in understanding the research landscape, identifying opportunities for improvement, and promoting significant advancements in the understanding and treatment of this medical condition.

**Methods:** An observational, descriptive, and cross-sectional study was conducted through a bibliometric analysis of the scientific output of the Familial Hypophosphatemic Rickets published in journals indexed in Scopus during 2020-2022. To retrieve the publications, Scopus was accessed on April 4, 2023, and an advanced search was performed using a filter by title, abstract and key words, source (journals), publication year, and type of article (article and review). The search terms used were extracted from the PubMed Medical Subject Headings (MeSH) related to the disease included in the MeSH catalog. Additionally, an analysis of co-occurrence between countries and keywords was carried out with VOSviewer software.

**Results:** This study identified 1,269 articles on hypophosphatemic rickets (938 articles and 331 reviews). In total, 39,548 citations were received, with an H index of 95. The majority of the articles (76.9%) were published in high-impact journals (Q1 and Q2 journals). Scientific production has shown a growing trend in recent years. The countries with the highest scientific production are the U.Ss, Japan, and the United Kingdom, considering that middle- and low-income countries contribute less to international scientific production.

**Conclusions:** Scientific production has shown sustained growth in recent years. The U.Ss solidifies itself as the country leading scientific production on hypophosphatemic rickets.

## 1. Background

Hypophosphatemic rickets comprise a group of genetic disorders characterized by renal phosphate wasting, resulting in hypophosphatemia, rickets, and normal serum calcium levels. The clinical features of these patients include slow growth/short stature, bone pain, and skeletal deformities. Hypophosphatemic rickets typically present in infancy or early childhood with skeletal deformities and growth plate abnormalities (1). The most common cause of hypophosphatemic rickets is genetic factors, such as X-linked hypophosphatemia (XLH) (2)

An estimated prevalence of less than 1 case per 1,000,000 people has been reported for autosomal dominant hypophosphatemic rickets and hypophosphatemic rickets with hypercalciuria. (3,4). In some regions, the estimated incidence reaches up to 2.03 cases per 100,000 inhabitants (5). The worldwide incidence of XLH is reported to be 1 to 9 cases per 1,000,000 habitants (6).

XLH has been described as a disabling condition for those affected, with a negative impact on physical functioning, activities of daily living, mental health, social life, and leisure activities. In adulthood, it is associated with a substantial disease burden and decreased quality of life, where individuals often experience severe pain and progressive disability (7–10).

Bibliometrics is the application of mathematics and statistical methods to any written source based on communication facets and considers elements such as authors, publication title, document type, language, abstract, and keywords or descriptors (11,12). Bibliometric studies, in any branch of science, represent a necessary and unavoidable reality in the era of information and communications. They serve not only as an instrument for evaluating the scientific production of a particular subject but also as a means to enhance and elevate the excellence of scientific research to higher levels (12).

Research in the health sciences serves as the cornerstone upon which health services develop and progress, refining medical care at various levels. Evaluating scientific production on a specific disease or health issue is strategically important for health policy decision-making at the national or regional level. This provides a general understanding of the importance attributed by the scientific community to a topic based on the research generated around it. The evaluation of the scientific production on familial hypophosphatemic rickets aids in understanding the research landscape, identifying opportunities for improvement, and promoting significant advancements in the understanding and treatment of this medical condition.

As far as has been reviewed, no studies evaluating the scientific production of familial hypophosphatemic rickets have been found in the main databases for scientific information retrieval. For this reason, the primary objective of this work is to characterize the scientific production of original articles and review articles on familial hypophosphatemic rickets in indexed journals in Scopus. This includes examining collaborations in these publications and their impact in terms of study citations globally. Similarly, trends in related research are presented.

## 2. Methods

### 2.1 Design

An observational, descriptive, and cross-sectional study was conducted through a bibliometric analysis of the scientific output of Familial Hypophosphatemic Rickets published in journals indexed in Scopus.

### 2.2 Bibliometric indicators

The following bibliometric indicators were studied:

- Number of documents (Ndoc).
- Articles in English (Ndoc Eng). Articles published in English.
- Non-English article (Ndoc Non-Eng). Articles published in a language other than English.
- Overlap (Ndoc Overlap). Articles published in two languages.
- Citations (NCit). Total citations received from articles indexed in Scopus.
- Cited articles (Cited doc). Total number of published articles that have been cited at least once according to Scopus.
- Citations per document (Cpd). Average number of received citations.
- H-index. This index considers both the number of articles and the citations they receive. An author has an h = x index if he/she has x articles that have been cited at least x times (13). This indicator is also used to characterize groups (a group of authors, a department, or a country).
- R-index: Square root of the total number of citations received by the core articles contributing to the h-index (14).
- Growth rate (GR): percent change in the number of articles published in a domain with respect to the previous year, calculated as follows: GRn = [(Ndocn−Ndocn−1)/Ndocn−1] * 100, where **n** is the year (15).
- Quartiles (Q). According to the SCImago Journal & Country Rank (SJR), the journals indexed in Scopus are placed in quartiles, where those in the first quartile have the highest impact. There are journals that do not appear in the ranking (nonranked) due to their recent inclusion in the database.
- High-quality publications (Ndoc Q1). Percentage of publications in journals included in the quartile of maximum visibility.
- Scientific leadership (% Lead). Percentage of articles from a country in which the corresponding author belongs to a national institution. These are referred to as lead documents.
- % Q1 Lead (Ndoc Q1 Lead). The percentage of articles in journals included in the first quartile in which the corresponding author is affiliated with an institution who belongs to the same country.

### 2.3 Data collection and search strategy

To retrieve the publications, Scopus (Elsevier BV Company, Netherlands, http://www.scopus.com) was the database chosen to identify the studies because it is considered the most complete database worldwide, encompassing 100% of the publications indexed in Medline. Scopus was accessed on April 4, 2023, and an advanced search was performed using a filter by title, abstract and key words, source (journals), publication year, and type of article (article and review). The search terms used were extracted from the PubMed Medical Subject Headings (MeSH) related to the disease included in the MeSH catalog. The **search strategy** was as follows:

TITLE-ABS-KEY ( “x linked hypophosphatemic” OR “x-linked hypophosphatemic” OR “Generalized Resistance To 1,25 Dihydroxyvitamin D” OR “Generalized Resistance To 1,25-Dihydroxyvitamin D” OR “Hereditary Hypophosphatemic Rickets”

OR “Hereditary Vitamin D Resistant Rickets” OR “Hereditary Vitamin D-Resistant Rickets” OR “Hypocalcemic Vitamin D Resistant Rickets” OR “Hypocalcemic Vitamin D-Resistant Rickets” OR “X Linked Hypophosphatemia” OR “X-Linked Hypophosphatemia” OR “Familial Hypophosphatemic Rickets” OR “Hereditary Hypophosphatemic Rickets” OR “X Linked Dominant Hypophosphatemic Rickets” OR “X Linked Recessive Hypophosphatemic Rickets” OR “X-Linked Recessive Hypophosphatemic Rickets” OR “X-Linked Dominant Hypophosphatemic Rickets” OR “Familial Hypophosphatemic Rickets” OR “Hereditary Hypophosphatemic Rickets” OR “Hereditary Vitamin D Resistant Rickets” OR “Hereditary Vitamin D-Resistant Rickets” OR “X-Linked Hypophosphatemic Rickets” OR “Vitamin D Resistant Rickets With End Organ Unresponsiveness To 1,25 Dihydroxycholecalciferol” OR “Hereditary Vitamin D Resistant Rickets” OR “X Linked Vitamin D Resistant Rickets” OR “X-Linked Vitamin D Resistant Rickets” OR “Vitamin D-Resistant Rickets With End-Organ Unresponsiveness To 1,25-Dihydroxycholecalciferol” OR “Hereditary Vitamin D-Resistant Rickets” OR “X-Linked Vitamin D-Resistant Rickets” OR “X Linked Hypophosphatemia” OR “X-Linked Hypophosphatemia” OR “X-Linked Hypophosphatemic Rickets” ) AND SRCTYPE ( j ) AND PUBYEAR > 1999 AND ( LIMIT-TO ( DOCTYPE , “ar” ) OR LIMIT-TO ( DOCTYPE , “re” ) ) AND ( EXCLUDE ( PUBYEAR , 2023 ) )

Visualization map analysis according to the authors’ keywords. was created using VOSviewer version 1.6.15 to analyze the co-occurrence of the collaborating countries and keywords. The graphical interpretation is based on the grouping guidelines (keywords) and the distance between countries (co-occurrence networks). Additionally, descriptive statistics were performed using Microsoft Excel 2019®, calculating the absolute and relative frequencies for each variable of the study.

### 2.4 Ethics

The data were downloaded from public databases; therefore, no ethical approval was obtained.

## 3. Results

### 3.1 General bibliometric indicators

Initially, 1309 articles were retrieved, and after normalization, 1269 were identified. Scopus indices 1269 articles that received 39548 citations, with an average of 31.2 citations per document (Table 1). The ratio of original articles to reviews was 2.8. The overall values of the H and R indices are 95 and 143.2, respectively. Additionally, 86% of the publications have been cited (n = 1091); 1006 have received from 1 to 100 citations; 56 articles, from 101 to 200 citations; 14, from 201 to 300 citations; four, from 301 to 400 citations; five, from 401 to 500 citations; and the remaining six, more than 500. The most cited article has received 1284 citations and was published in *Nature Genetics* journal, which belongs to the first quartile of visibility in Scopus titled “Autosomal dominant hypophosphataemic rickets is associated with mutations in FGF23” by Kenneth E. White and Bettina Lorenz-Depiereux et al. (**Supplementary material**).

**Table 1.** General bibliometric indicators of the world scientific output.

| Indicator | Value |
| --- | --- |
| Ndoc | 1269 |
| Ndoc Eng (%) | 1158 (91.3) |
| Ndoc Non-Eng (%) | 96 (7.7) |
| Ndoc Overlap (%) | 15 (1.2) |
| Articles (%) | 938 (73.9) |
| Reviews (%) | 331 (26.1) |
| NCit | 39548 |
| Cited doc (%) | 1091 (86.0) |
| Cpd | 31.2 |
| H index | 95 |
| R index | 143.2 |

More than half of the articles (56.6%; n=718) were published in high-visibility journals (Q1), all in the English language, with 70.9% of them corresponding to original articles. Among these, 675 documents were cited, with an average H-index of 91 (Table 2).

**Table 2.** Bibliometric indicators according to quartiles of visibility.

| Indicators | Q1 | Q2 | Q3 | Q4 | Nonranked |
| --- | --- | --- | --- | --- | --- |
| Ndoc (%) | 718 (56.6) | 257 (20.3) | 117 (9.2) | 79 (6.2) | 98 (7.7) |
| Ndoc Eng (%) | 718 (100) | 253 (98.4) | 96 (82.1) | 36 (45.6) | 55 (56.1) |
| Ndoc Non-Eng (%) | - | 2 (0.8) | 11 (9.4) | 41 (51.9) | 42 (42.9) |
| Ndoc Overlap (%) | - | 2 (0.8) | 10 (8.5) | 2 (2.5) | 1 (1.0) |
| Articles (%) | 538 (74.9) | 204 (79.4) | 83 (70.9) | 59 (74.7) | 54 (55.1) |
| Reviews (%) | 180 (25.1) | 53 (20.6) | 34 (29.1) | 20 (25.3) | 44 (44.9) |
| NCit | 33157 | 3764 | 949 | 244 | 1434 |
| Cpd | 46.2 | 14.6 | 11.1 | 3.1 | 14.6 |
| Cited doc (%) | 675 (94.0) | 215 (83.7) | 93 (79.5) | 38 (48.1) | 70 (71.4) |
| H index | 91 | 34 | 17 | 7 | 17 |
| R index | 140.1 | 44.8 | 24.4 | 13.2 | 34.6 |

Table 3 displays the scientific production over the years, along with corresponding bibliometric indicators. Overall, there is a noticeable trend of growth in the scientific production of hypophosphatemic rickets. Starting in 2020, more than 100 publications were indexed in Scopus.

**Table 3.** Annual bibliometric indicators of the world scientific output.

### 3.2 Bibliometric indicators of output and impact on journals

The bibliometric indicators and impact on journals with ≥15 papers are shown in Table 4. The *Journal of Bone and Mineral Research* is the journal most frequently chosen by researchers for publication. Its 67 articles have received 4507 citations and have an H-index of 31. In terms of impact, as evaluated through the citations received, the articles published in this journal had the highest results, reaching a high average number of citations per paper (67.3).

**Table 4.** Bibliometric indicators of output and impact on journals with fifteen or more articles.

### 3.3 Scientific leadership

Table 5 and Figure 1 show the distribution of scientific production by country with fifty or more articles, which shows a predominance of documents from the U.S. (n = 454), representing 35.7% of the total number of published articles. In terms of impact according to the number of citations received, this figure far surpasses that of the other countries since its articles have received 24606 citations, with a high average number of citations per document and H and R indices (Cpd = 54.2; H index = 80; R index = 156.9). Japan is the country with the second highest scientific production since its 160 articles have received 6388 citations (Cpd = 69.29; H index = 42; R index = 412.94).

**Figure 1.**
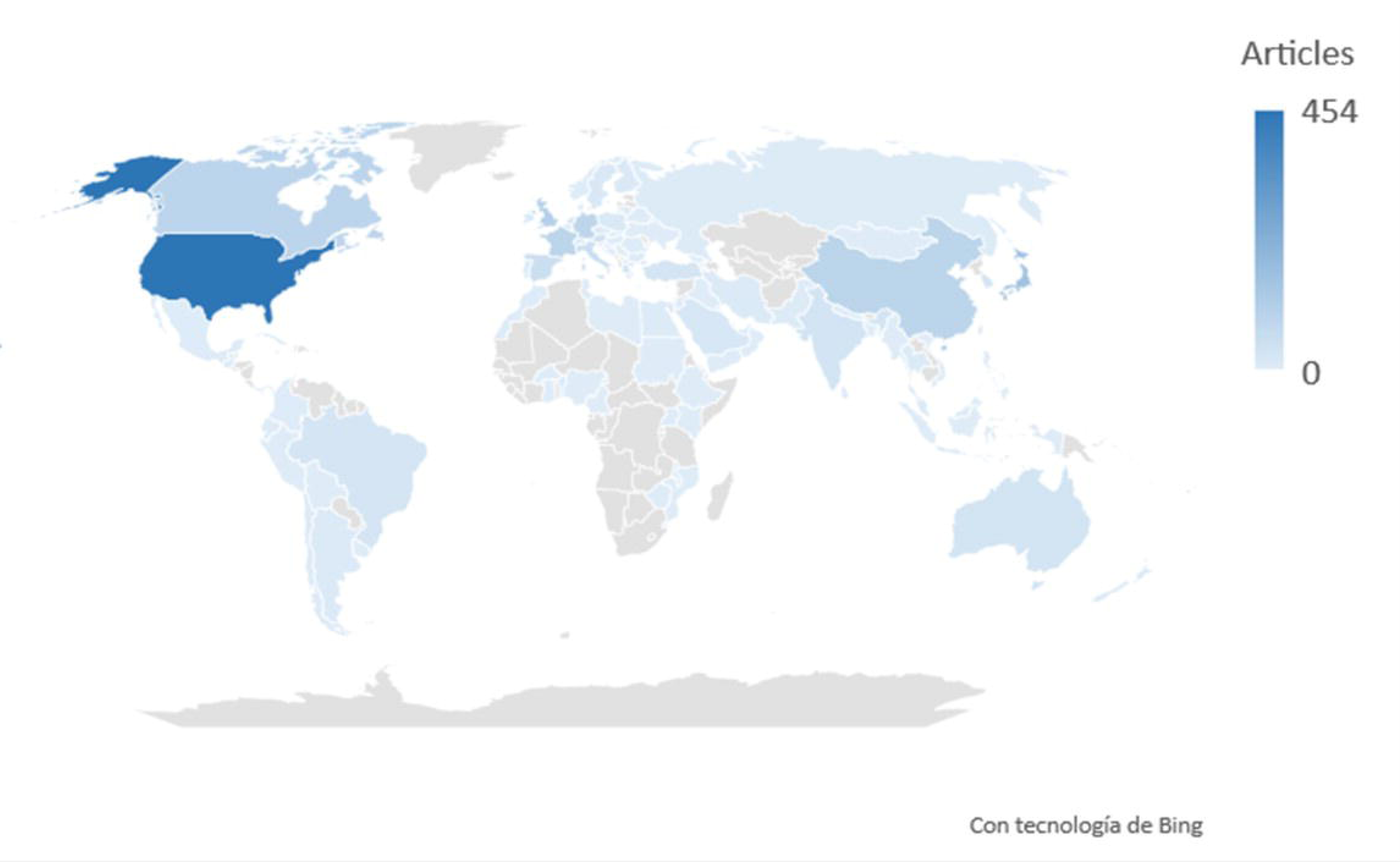
Scientific production by country.

**Table 5.**
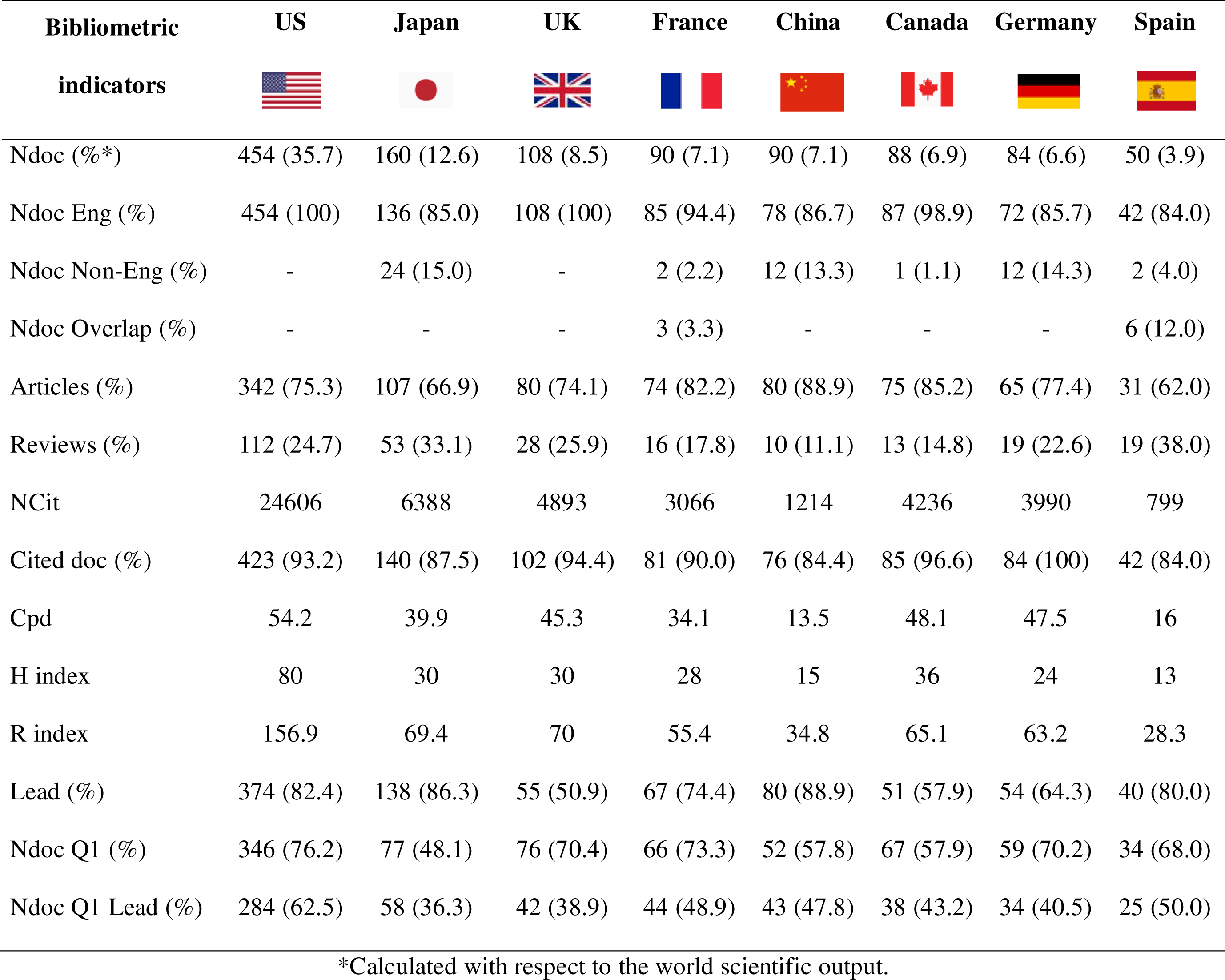
Bibliometric indicators of the output, impact, and scientific leadership of countries with fifty or more articles.

| Bibliometric<br>indicators | US | Japan | UK | France | China | Canada | Germany | Spain |
| --- | --- | --- | --- | --- | --- | --- | --- | --- |
| Ndoc (%*) | 454 (35.7) | 160 (12.6) | 108 (8.5) | 90 (7.1) | 90 (7.1) | 88 (6.9) | 84 (6.6) | 50 (3.9) |
| Ndoc Eng (%) | 454 (100) | 136 (85.0) | 108 (100) | 85 (94.4) | 78 (86.7) | 87 (98.9) | 72 (85.7) | 42 (84.0) |
| Ndoc Non-Eng (%) | - | 24 (15.0) | - | 2 (2.2) | 12 (13.3) | 1 (1.1) | 12 (14.3) | 2 (4.0) |
| Ndoc Overlap (%) | - | - | - | 3 (3.3) | - | - | - | 6 (12.0) |
| Articles (%) | 342 (75.3) | 107 (66.9) | 80 (74.1) | 74 (82.2) | 80 (88.9) | 75 (85.2) | 65 (77.4) | 31 (62.0) |
| Reviews (%) | 112 (24.7) | 53 (33.1) | 28 (25.9) | 16 (17.8) | 10 (11.1) | 13 (14.8) | 19 (22.6) | 19 (38.0) |
| NCit | 24606 | 6388 | 4893 | 3066 | 1214 | 4236 | 3990 | 799 |
| Cited doc (%) | 423 (93.2) | 140 (87.5) | 102 (94.4) | 81 (90.0) | 76 (84.4) | 85 (96.6) | 84 (100) | 42 (84.0) |
| Cpd | 54.2 | 39.9 | 45.3 | 34.1 | 13.5 | 48.1 | 47.5 | 16 |
| H index | 80 | 30 | 30 | 28 | 15 | 36 | 24 | 13 |
| R index | 156.9 | 69.4 | 70 | 55.4 | 34.8 | 65.1 | 63.2 | 28.3 |
| Lead (%) | 374 (82.4) | 138 (86.3) | 55 (50.9) | 67 (74.4) | 80 (88.9) | 51 (57.9) | 54 (64.3) | 40 (80.0) |
| Ndoc Q1 (%) | 346 (76.2) | 77 (48.1) | 76 (70.4) | 66 (73.3) | 52 (57.8) | 67 (57.9) | 59 (70.2) | 34 (68.0) |
| Ndoc Q1 Lead (%) | 284 (62.5) | 58 (36.3) | 42 (38.9) | 44 (48.9) | 43 (47.8) | 38 (43.2) | 34 (40.5) | 25 (50.0) |

\*Calculated with respect to the world scientific output.

The U.Ss is the country with the highest leadership in terms of scientific production, where 82.4% of its articles have a corresponding author affiliated with a national institution. Of these articles, 62.5% were published in first quartile journals.

The most productive institutions were Indiana University School of Medicine (n=64), Université McGill (n=50), Yale School of Medicine (n=50), AP-HP Assistance Publique - Hopitaux de Paris (n=49), Université Paris Cité (n=41), Inserm (n=40) and Harvard Medical School (n=40).

There is extensive international scientific collaboration led by researchers in the U.Ss, Japan and Europe (Figure 2). The middle- and low-income countries, fundamentally in Africa and Latin America, are the least represented and productive in relation to research on familiar hypophosphatemic rickets.

**Figure 2.**
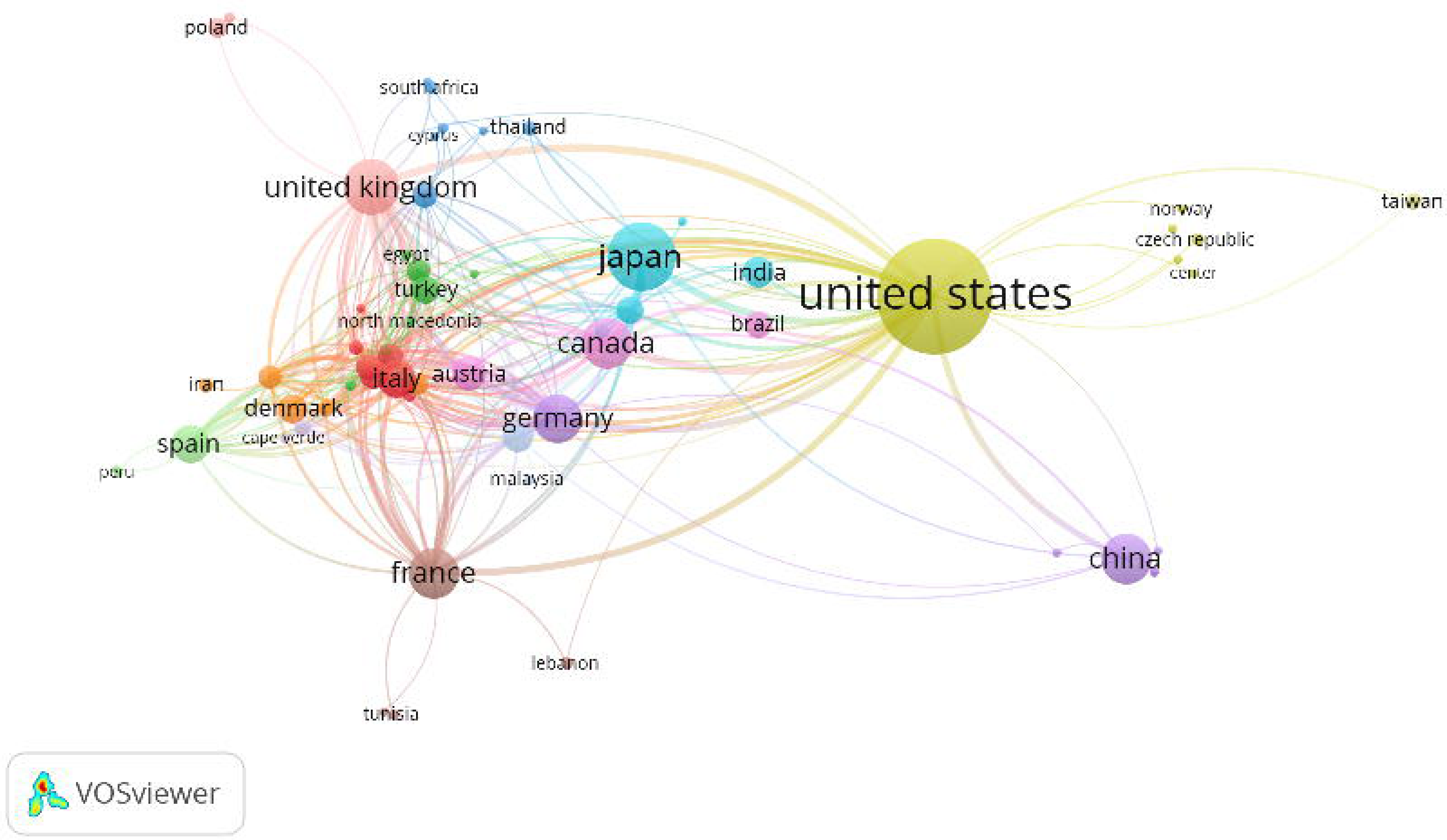
Scientific collaboration networks among countries with ≥5 articles published (Scopus, 2000–2022).

### 3.5 Research trends

Of the 1862 authors’ keywords, 108 were selected for analysis in VOSviewer and were chosen based on their occurrence in at least 6 documents (Figure 3). The most frequently used keywords by the authors were rickets (n=189), FGF23 (n=122), hypophosphataemia (n=137), PHEX (n=117), phosphate (n=84), osteomalacia (n=81), vitamin D (n=81), X-linked hypophosphatemic rickets (n=74) and burosumab (n=59).

**Figure 3.**
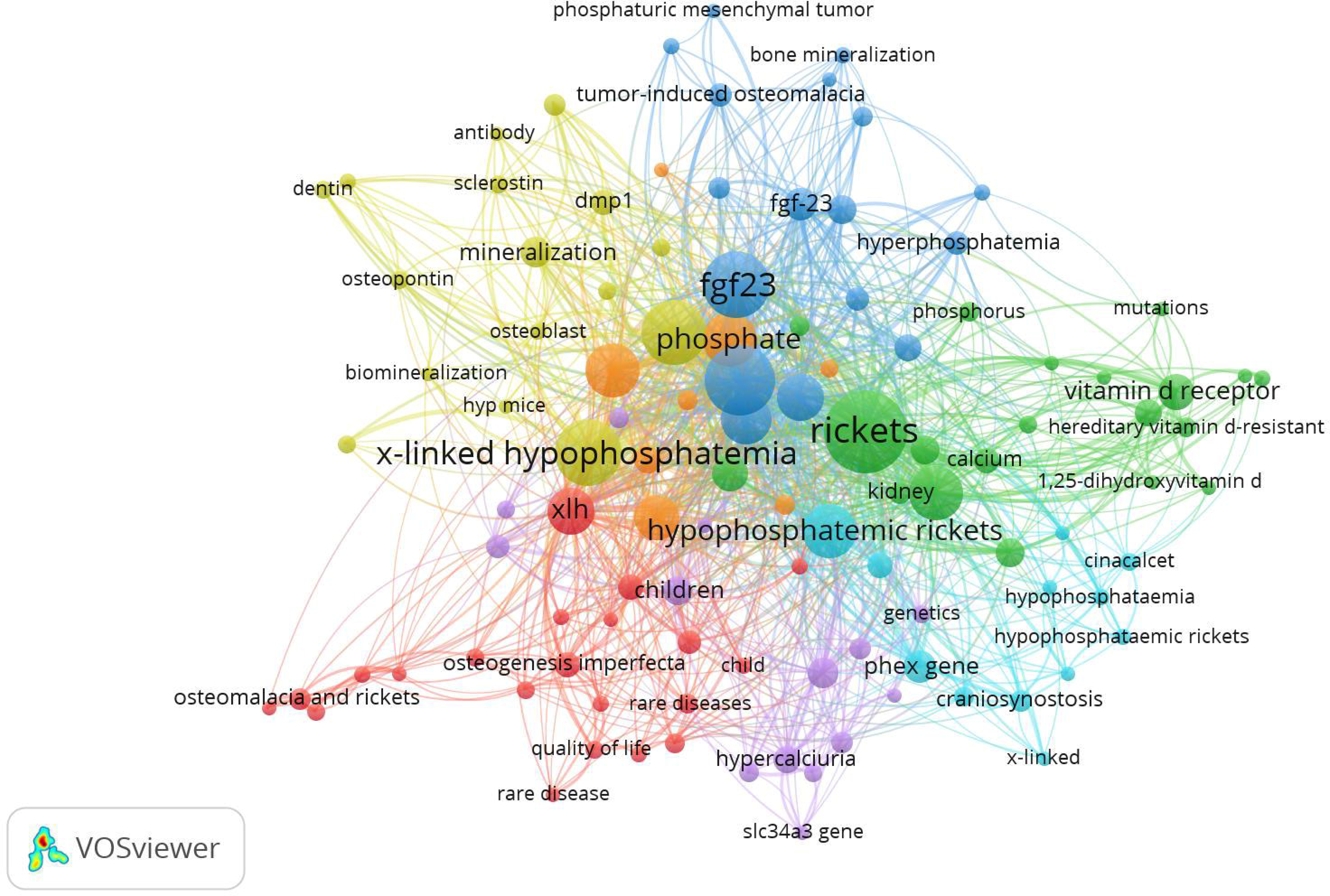
Author keywords and research topics. The map is based on text data from the scientific production of XLH (2000–2022).

The identification of the 108 authors’ keywords was divided into five different groups, where the following terms were used for each: 1) “genetic implications” (PHEX, mutation, Hyp mice/Hyp mouse); 2) “management and treatment” (burosumab, phosphate, cinacalcet); 3) “pophysiology relationships and protein related” (Dmp-1, osteopontin, sclerostin, 1,25 dihydroxyvitamin D, phosphate metabolism); 4) “complication and quality of life” (nontrocalcinosis, hyperparathyroidism); and 5) “clinical and analytical manifestations relationship” (hypophosphatemia, Rickets, Osteomalacia, phosphaturia). There was no apparent association between color and research theme, which may be due to the overlap of these terms across different articles.

Figure 4 shows the evolution of keywords and research trends in familial hypophosphatemic rickets over the years. In the last 5 years, research on the quality of life and chronic complications of this disease has gained prominence, becoming a trend in the investigation of hypophosphatemic rickets and XLH.

**Figure 4.**
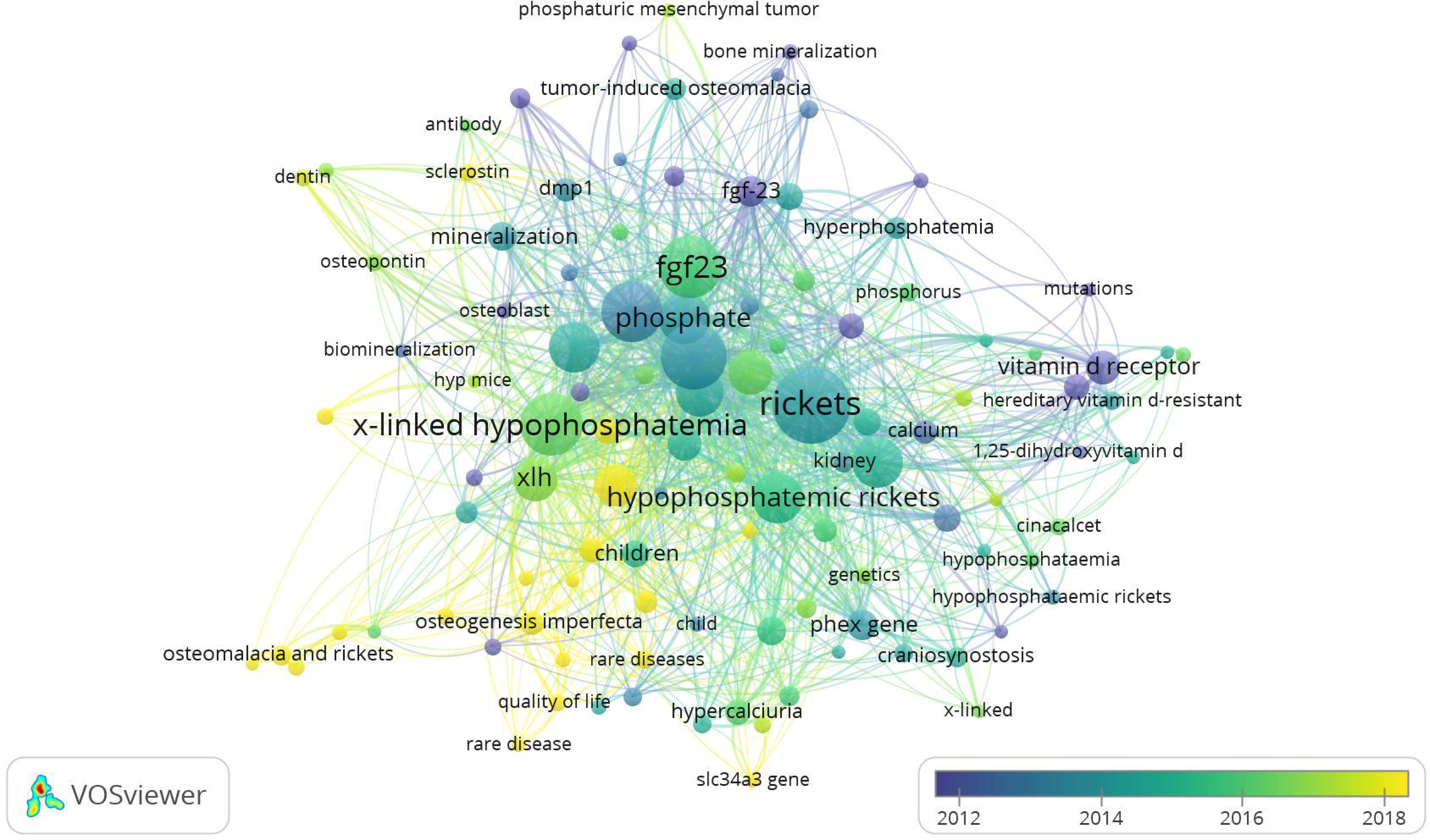
Year’s authors keywords and research topics. The map is based on text data from the scientific production of XLH (2000–2022).

## 4. Discussion

The scientific landscape of rare diseases has rapidly evolved, with expectations for this change to accelerate. Individuals living with rare diseases increasingly benefit from new therapies, and related research has been conducted (14). By definition, rare diseases are uncommon, leading to fewer patients, physicians, and researchers dedicated to them, as is the case with familial hypophosphatemic rickets. The infrastructure required to conduct studies in this area can at times become a costly endeavor, especially when considering per-participant research expenses.

The present research constitutes the first study analyzing the individual scientific basis of a rare disease, specifically familial hypophosphatemic rickets. In comparison to other medical conditions, scientific production on familial hypophosphatemic rickets could be considered low, based on the finding of only 1269 indexed documents over 22 years. Given its status as a group of rare and low-prevalence diseases, scientific activity focused on this disease is reserved for specialized research groups and reference institutions in the field. The prevalence of other diseases, whose diagnosis is centered around the pediatric age, is greater than that of other diseases (16–19), although in some of these studies, the evaluated period was longer than that of our study.

A higher number of publications in English is an expected outcome. English has become the dominant language for global scientific communication. In other studies in which scientific production and communication language have been evaluated in specific areas of pediatrics, English was also determined to be the most widely used language for writing and publishing scientific articles (17,18,20–22). Several factors contribute to the widespread use of English in scientific publications, including global collaboration (where researchers from different countries and linguistic backgrounds collaborate and communicate effectively), accessibility (English publications are more accessible to a global audience), and standardization (the use of a common language helps standardize scientific terminology and facilitates clear communication of complex concepts). Furthermore, many prestigious scientific journals, especially those with high impact factors, are published in English. Researchers often target these journals to enhance the visibility and impact of their work, as measured by citation metrics. Some journals and conferences offer language support or translations to accommodate researchers who do not have English as their native language. However, it is likely that the trend of using English as the primary language for identifying scientific publications will continue in the foreseeable future.

The most productive publications on familial hypophosphatemic rickets are expected to be found in the SJR thematic area corresponding to *Endocrinology, Diabetes, and Metabolism*. The *Journal of Bone and Mineral Research*, *Bone* and *Journal of Clinical Endocrinology and Metabolism* are the most productive journals in this field, with the highest number of documents and citations and the highest H-index. Another author also found the Journal of Clinical Endocrinology and Metabolism to be one of the most cited journals on the scientific productivity of Spanish pediatrics (21). A study conducted by Chhapola V., et al. (20), where they analyzed citations of classic pediatric studies, determined that the pediatric subspecialties of growth and development and endocrinology were the most cited and productive.

With regard to the growth rate of scientific production, it can be asserted that it has fluctuated since 2000, with both positive and negative values. However, since 2017, it has remained consistently positive, reaching its peak value in 2022 at 11.2. Notably, before 2020, no more than 100 publications on familial hypophosphatemic rickets were indexed in the database. In recent years, this figure has only marginally surpassed 100 points. Therefore, in comparison with other thematic areas (e.g., COVID-19 (15), diabetes (16), or congenital heart disease (17)), scientific production in familial hypophosphatemic rickets is relatively limited. It is important to consider that these conditions have low incidence and prevalence rates and are sometimes underdiagnosed.

Globally, in correspondence with other research areas (16–18,20,23), the U.S is the most productive country in terms of familial hypophosphatemic rickets, contributing to more than a quarter of the world’s scientific production, and 60% of its articles are published. This fact may be attributed, among other factors, to the presence of some of the world’s leading medical research centers and specialized hospitals for genetic diseases in the U.S. Significant investment in research and development in the medical and scientific fields by government agencies, nonprofit organizations, and the private industry contributes to research leadership. These investments enable the conduct of studies, clinical trials, and the development of innovative treatments. The advanced technological infrastructure in the U.Ss, which includes genetic sequencing facilities, bioinformatics, and other cutting-edge technologies, allows researchers to conduct detailed studies on genetic diseases and develop more effective therapeutic approaches. The quality of education and the ability to attract and retain highly trained professionals in scientific and medical fields are crucial factors for research leadership. In contrast, according to the bibliometric analysis by Yuan T., et al. (24), China has emerged as the most productive country for genetic research on pediatric cataracts.

International collaboration networks are crucial in medical research. Research collaboration can help increase scientific productivity (25). The U.S leads in terms of its broad collaboration network, followed by Japan, which is the second most productive country. Third, the European region as a whole, led by the United Kingdom, France, Germany, and Spain, could be attributed. Several of the primary research groups are located in these countries. These results align with the findings of other authors, where the U.S. and Europe led scientific collaboration (16,17). Low-income countries, primarily in Africa and some in Latin America, have the lowest scientific production of XLH, and in some cases, it is nonexistent. Several bibliometric studies in the field of pediatrics have shown low indexed scientific production in these regions (17,18,23).

The International Rare Disease Research Consortium (IRDiRC) has established a promising initiative in rare disease research; nevertheless, there is still much work to be done in this field. This is especially pertinent given that less than 6% of rare diseases have approved treatments, and the majority of drug development efforts are concentrated on a limited number of diseases (19). Precise data on the quantification and progress of XLH are not currently available. The International X-Linked Hypophosphatemia (XLH) Registry provides an approach to these data (26). Other projects and research groups, both basic and clinical, have focused on the study of familial hypophosphatemic rickets and other rare diseases, with primary tubulopathy as the physiopathological basis (27). There is a need to develop a common high-quality platform for registering rare bone and mineral conditions (28).

The analysis of keywords revealed several significant trends in research on X-linked hypophosphatemic rickets (XLH). The most frequent keywords provide a comprehensive view of the focus areas in the scientific literature. The generic name of the disease, “Rickets,” and “X-linked hypophosphatemic rickets” are key terms, emphasizing the importance of understanding the disease from both clinical and genetic perspectives. Terms such as “FGF23,” “PHEX,” and “phosphate” suggest a focus on understanding the underlying pathophysiological and genetic mechanisms of XLH. The presence of “Osteomalacia” as a keyword indicates an emphasis on investigating complications associated with the disease, delving into long-term effects on bone tissue.

With the introduction of burosumab as a new therapy for XLH (29), research on this topic has influenced its positioning as a keyword used by authors in both basic research and clinical trials. The inclusion of “burosumab” and “vitamin D” reflects the interest in therapeutically addressing XLH, with special attention given to the new therapy burosumab. The use of burosumab as a keyword indicates its significant impact on research, both in basic studies and clinical trials. The focus on “chronic complications” suggested a shift toward understanding the long-term effects of the disease. Additionally, the study of “quality of life” reflects a growing interest in addressing broader aspects of patient well-being. The evolution of keywords reflects not only progress in the scientific and therapeutic understanding of X-linked hypophosphatemic rickets but also a shift toward more holistic considerations that encompass the quality of life and chronic complications of patients.

Some limitations of our study should be addressed in future scientometric research. Despite selecting Scopus for its recognition as the most comprehensive database, it is possible that several studies published in nonindexed journals, those in the process of indexing, or preprint repositories may have been excluded from the analysis. Additionally, many studies from low-income countries tend to be published in local journals that are not indexed, potentially affecting their perceived productivity in the context of hypophosphatemic rickets. Moreover, due to constantly changing citation volumes over time, the results of our study are temporary and valid for the present study. Nevertheless, we believe our study provides a detailed international scientometric analysis of the scientific production of familial hypophosphatemic rickets.

The results suggest the need to promote international cooperation through engagement with leading research groups and institutions.

## Supporting information

Table 3. Annual bibliometric indicators of the world scientific output.

Table 4. Bibliometric indicators of output and impact on journals with fifteen or more articles.

## Data Availability

Harvard Dataverse https://doi.org/10.7910/DVN/X0NUZY

## Funding

This research was funded by the Research Vice-Rectorate of the University of Oviedo through the University of Oviedo Research Support and Promotion Plan (reference PAPI-22-PF-4).

### CRediT authorship contribution statement

Conceptualization: FHG, IECR. Data curation: FHG, IECR. Formal analysis: FHG, IECR. Funding acquisition: FHG, HGP, JRS, JMLG. Investigation: FHG, IECR. Methodology: FHG, IECR. Resources: FHG. Software: FHG, IECR. Supervision: HGP, JRS, JMLG, RFP, and IECR. Validation: FHG, IECR. Visualization: FHG, IECR. Writing – original draft: FHG. Writing – review and editing: FHG, HGP, JRS, JMLG, POC, RFP, IECR. All the authors have read and agreed to the published version of the manuscript.

### Availability of data and materials

The datasets generated and analyzed during the current study are available in the Harvard Dataverse repository https://doi.org/10.7910/DVN/X0NUZY.

### Declaration of competing interest

The authors declare no competing interests.

## Notes

### Competing Interest Statement

The authors have declared no competing interest.

## REFERENCES

1. Fuente R, Gil-Peña H, Claramunt-Taberner D, Hernández O, Fernández-Iglesias A, Alonso-Durán L, et al. X-linked hypophosphatemia and growth. Vol. 18, Reviews in Endocrine and Metabolic Disorders. Springer New York LLC; 2017. p. 107–15. 10.1007/s11154-017-9408-1

2. Ackah SA, Imel EA. Approach to Hypophosphatemic Rickets. J Clin Endocrinol Metab. 2022 Dec 17;108(1):209–20. 10.1210/clinem/dgac488

3. Orphanet. Orphanet Portal de información de enfermedades raras y medicamentos huérfanos . 2017. Raquitismo hipofosfatemico autosomico dominante. https://www.orpha.net/consor/cgi-bin/OC_Exp.php?lng=ES&Expert=89937

4. Orphanet. Orphanet Portal de información de enfermedades raras y medicamentos huérfanos . 2017. Raquitismo hipofosfatémico hereditario con hipercalciuria. https://www.orpha.net/consor/cgi-bin/OC_Exp.php?Expert=157215&lng=ES

5. Huertas-Quintero JA, Losada-Trujillo N, Cuellar-Ortiz DA, Velasco-Parra HM. Hypophosphatemic Rickets in Colombia: A Prevalence-Estimation Model in Rare Diseases. 2018. The Lancet Regional Health - Americas. 2022 Mar;7:100131. 10.1016/j.lana.2021.100131

6. Orphanet. Portal de información de enfermedades raras y medicamentos huérfanos. 2017. Hipofosfatemia ligada al X. https://www.orpha.net/consor/cgi-bin/OC_Exp.php?Expert=89936&lng=ES

7. Seefried L, Smyth M, Keen R, Harvengt P. Burden of disease associated with X-linked hypophosphataemia in adults: a systematic literature review. Vol. 32, Osteoporosis International. Springer Science and Business Media Deutschland GmbH; 2021. p. 7–22. 10.1007/s00198-020-05548-0

8. Yanes MIL, Diaz-Curiel M, Peris P, Vicente C, Marin S, Ramon-Krauel M, et al. Health-related quality of life of X-linked hypophosphatemia in Spain. Orphanet J Rare Dis. 2022 Dec 29;17(1):298. 10.1186/s13023-022-02452-0

9. Jandhyala R. Concordance between the schedule for the evaluation of individual quality of life-direct weighting (SEIQoL-DW) and the EuroQoL-5D (EQ-5D) measures of quality of life outcomes in adults with X-linked hypophosphatemia. Orphanet J Rare Dis. 2022 Dec 23;17(1):81. 10.1186/s13023-022-02250-8

10. Rodríguez-Rubio E, Gil-Peña H, Chocron S, Madariaga L, de la Cerda-Ojeda F, Fernández-Fernández M, et al. Phenotypic characterization of X-linked hypophosphatemia in pediatric Spanish population. Orphanet J Rare Dis. 2021 Dec 1;16(1). 10.1186/s13023-021-01729-0

11. Gorbea Portal S. Una nueva perspectiva teórica de la bibliometría basada en su dimensión histórica y sus referentes temporales. Investigación Bibliotecológica: Archivonomía, Bibliotecología e Información. 2016 Sep;30(70):11–6. 10.1016/j.ibbai.2016.10.001.

12. Solano Lopez E, Castellanos Quintero S, Lopez Rodriguez Del Rey M, Hernandez Fernandez J. La bibliometría: una herramienta eficaz para evaluar la actividad científica postgraduada. MediSur. 2009;7(4):59–62. http://scielo.sld.cu/scielo.php?script=sci_arttext&pid=S1727-897X2009000400011&lng=es&nrm=iso

13. Hirsch JE. An index to quantify an individual’s scientific research output. Proceedings of the National Academy of Sciences. 2005 Nov 15;102(46):16569–72. 10.1073/pnas.0507655102

14. Jin BH, Liang LM, Rousseau R, Egghe L. The R- and AR-indices: Complementing the h-index. Chinese Science Bulletin. 2007 Mar;52(6):855–63. 10.1007/s11434-007-0145-9

15. Zacca-González G. Producción científica latinoamericana en Salud Pública: Cuba en el contexto regional, Scopus 2003-2011 [Internet]. 2015 [cited 2023 Nov 26]. Available from: http://hdl.handle.net/10481/40902

16. Martynov I, Klima-Frysch J, Schoenberger J. A scientometric analysis of neuroblastoma research. BMC Cancer. 2020 May 29;20(1). 10.1186/s12885-020-06974-3

17. Friedmacher F, Pakarinen MP, Rintala RJ. Congenital diaphragmatic hernia: a scientometric analysis of the global research activity and collaborative networks. Vol. 34, Pediatric Surgery International. Springer Verlag; 2018. p. 907–17. 10.1007/s00383-018-4304-7

18. Karabulut A, Kaya M. Crohn’s disease from past to present: Research trends and global outcomes with scientometric analysis during 1980 to 2022. Medicine (United States). 2023 Sep 1;102(35):E34817. 10.1097/MD.0000000000034817

19. Cortese S, Sabe M, Angriman M, Solmi M. The Italian contribution to pediatric sleep medicine: A scientometric analysis. Sleep Med. 2023 Jul 1;107:164–70. 10.1016/j.sleep.2023.05.002

20. Chhapola V, Tiwari S, Deepthi B, Kanwal SK. Citation classics in pediatrics: a bibliometric analysis. World Journal of Pediatrics. 2018 Dec 1;14(6):607–14. 10.1007/s12519-018-0146-6

21. Alonso-Arroyo A, González De Dios J, Bolanos-Pizarro M, Castelló-Cogollosc L, González-Alcaide G, Navarro-Molina C, et al. Analysis of the scientific productivity and impact of Spanish paediatrics (2006-2010). An Pediatr (Engl Ed). 2013;78(6). 10.1016/j.anpedi.2013.01.003

22. Corrales-Reyes IE, Hernández-García F, Mejia CR. COVID-19 and diabetes: Analysis of the scientific production indexed in Scopus. Diabetes and Metabolic Syndrome: Clinical Research and Reviews. 2021 May 1;15(3):765–70. 10.1016/j.dsx.2021.03.002

23. Zila-Velasque JP, Grados-Espinoza P, Cubas WS, Diaz-Barrera M, Pacheco-Mendoza J. Analysis of congenital heart disease research: Mapping impact, production and global collaboration. Heliyon. 2023 Aug 1;9(8). 10.1016/j.heliyon.2023.e19188

24. Tan Y, Jiang W, Hu LY, Shen YY, Chen H, Zou YS, et al. Hotspots and frontiers of genetic research on pediatric cataracts from 2013 to 2022: a scientometric analysis. Int J Ophthalmol. 2023;16(10):1682–91. 10.18240/ijo.2023.10.19

25. Shahmoradi L, Ramezani A, Atlasi R, Namazi N, Larijani B. Visualization of knowledge flow in interpersonal scientific collaboration network endocrinology and metabolism research institute. Vol. 20, Journal of Diabetes and Metabolic Disorders. Springer Science and Business Media Deutschland GmbH; 2021. p. 815–23. 10.1007/s40200-020-00644-8

26. Ariceta G, Beck-Nielsen SS, Boot AM, Brandi ML, Briot K, de Lucas Collantes C, et al. The International X-Linked Hypophosphatemia (XLH) Registry: first interim analysis of baseline demographic, genetic and clinical data. Orphanet J Rare Dis. 2023 Sep 27;18(1):304. 10.1186/s13023-023-02882-4

27. Mejía N, Santos F, Claverie-Martín F, García-Nieto V, Ariceta G, Castaño L. RenalTube: A network tool for clinical and genetic diagnosis of primary tubulopathies. Eur J Pediatr. 2013 Jun;172(6):775–80. 10.1007/s00431-013-1934-6

28. Priego Zurita AL, Grasemann C, Boarini M, Chapurlat R, Mordenti M, Javaid MK, et al. Data collection on rare bone and mineral conditions in Europe: The landscape of registries and databases. Eur J Med Genet. 2023 Dec;66(12):104868. 10.1016/j.ejmg.2023.104868

29. Sandy JL, Simm PJ, Biggin A, Rodda CP, Wall CL, Siafarikas A, et al. Clinical practice guidelines for paediatric X-linked hypophosphataemia in the era of burosumab. J Paediatr Child Health. 2022 May 1;58(5):762–8. 10.1111/jpc.15976

