## Supplementary material for "A Bibliometric Approach to Scientific Production on Familial Hypophosphatemic Rickets in Scopus (2000-2022)": Table 3. Annual bibliometric indicators of the world scientific output.

| **Year** | **Ndoc (%)** | **Growth rate (%)** | **Ndoc Eng (%)** | **Ndoc Non-Eng (%)** | **Ndoc Overlap (%)** | **Articles (%)** | **Reviews (%)** | **NCit** | **Cpd** | **Cited doc (%)** | **H index** | **R index** |
| --- | --- | --- | --- | --- | --- | --- | --- | --- | --- | --- | --- | --- |
| 2000 | 22 (1.7) | -4.3 | 21 (95.5) | 1 (4.5) | - | 19 (86.4) | 3 (13.6) | 1951 | 88.7 | 20 (90.9) | 15 | 43.8 |
| 2001 | 28 (2.2) | 27.3 | 26 (92.9) | 2 (7.1) | - | 23 (82.1) | 5 (17.9) | 1635 | 58.4 | 27 (96.4) | 16 | 39.8 |
| 2002 | 24 (1.9) | 14.3 | 24 (100) | - | - | 20 (83.3) | 4 (83.37) | 1642 | 68.4 | 24 (100) | 18 | 39.8 |
| 2003 | 15 (1.2) | -37.5 | 15 (100) | - | - | 14 (93.3) | 1 (6.7) | 2306 | 153.7 | 15 (100) | 13 | 47.8 |
| 2004 | 37 (2.9) | 146.6 | 31 (83.8) | 5 (13.5) | 1 (2.7) | 27 (73.0) | 10 (27.0) | 3159 | 85.4 | 34 (91.9) | 21 | 55.3 |
| 2005 | 33 (2.6) | -10.8 | 26 (78.8) | 7 (21.2) | - | 22 (66.7) | 11 (33.3) | 1181 | 35.8 | 28 (84.8) | 14 | 33.1 |
| 2006 | 34 (2.7) | 3.0 | 29 (85.3) | 5 (14.7) | - | 25 (73.5) | 9 (26.5) | 2409 | 70.9 | 32 (94.1) | 19 | 48.1 |
| 2007 | 34 (2.7) | - | 31 (91.2) | 3 (8.8) | - | 26 (76.5) | 8 (23.5) | 1567 | 46.1 | 32 (94.1) | 17 | 38.9 |
| 2008 | 48 (3.8) | 41.2 | 44 (91.7) | 3 (6.3) | 1 (2.1) | 34 (70.8) | 14 (29.2) | 2139 | 44.6 | 44 (91.7) | 21 | 44.1 |
| 2009 | 43 (3.4) | -10.4 | 36 (83.7) | 7 (16.3) | - | 26 (60.5) | 17 (39.5) | 1681 | 39.1 | 39 (90.7) | 21 | 39.8 |
| 2010 | 51 (4.0) | 18.6 | 42 (82.4) | 9 (17.6) | - | 35 (68.6) | 16 (31.4) | 2486 | 48.7 | 44 (86.3) | 23 | 48.2 |
| 2011 | 52 (4.1) | 2.0 | 48 (92.3) | 4 (7.7) | - | 38 (73.1) | 14 (26.9) | 1728 | 33.2 | 47 (90.4) | 24 | 38.8 |
| 2012 | 53 (4.2) | 1.9 | 47 (88.6) | 6 (11.3) | - | 30 (56.6) | 23 (43.4) | 1355 | 40.2 | 52 (98.1) | 24 | 46.2 |
| 2013 | 56 (4.4) | 5.7 | 47 (83.9) | 8 (15.1) | 1 (1.8) | 46 (82.1) | 10 (17.8) | 1575 | 28.1 | 48 (85.7) | 22 | 39.7 |
| 2014 | 62 (4.9) | 10.7 | 57 (91.9) | 4 (6.4) | 1 (1.6) | 47 (75.8) | 15 (24.2) | 1730 | 27.9 | 58 (93.5) | 21 | 41.6 |
| 2015 | 60 (4.7) | -3.2 | 55 (91.6) | 4 (6.6) | 1 (1.6) | 50 (83.3) | 10 (16.6) | 2199 | 36.7 | 56 (93.3) | 22 | 46.9 |
| 2016 | 57 (4.5) | -5.0 | 52 (91.2) | 4 (7) | 1 (1.7) | 46 (80.7) | 11 (19.3) | 1544 | 27.1 | 56 (98.2) | 22 | 39.3 |
| 2017 | 62 (4.9) | 8.8 | 58 (93.5) | 3 (4.8) | 1 (1.6) | 47 (75.8) | 15 (24.2) | 1111 | 17.9 | 56 (90.3) | 20 | 33.3 |
| 2018 | 68 (5.4) | 9.7 | 64 (94.1) | 4 (5.8) | - | 53 (77.9) | 15 (22.0) | 1788 | 26.3 | 61 (89.7) | 21 | 42.3 |
| 2019 | 74 (5.8) | 8.8 | 71 (95.9) | 3 (4.1) | - | 51 (68.9) | 23 (31.1) | 1518 | 20.5 | 67 (90.5) | 20 | 38.9 |
| 2020 | 111 (8.7) | 50 | 104 (93.7) | 5 (4.5) | 2 (1.8) | 79 (71.2) | 32 (28.8) | 1179 | 10.6 | 96 (86.5) | 17 | 34.3 |
| 2021 | 116 (9.1) | 4.5 | 105 (90.5) | 7 (6.0) | 4 (3.4) | 89 (76.7) | 27 (23.3) | 569 | 4.9 | 88 (75.8) | 11 | 23.9 |
| 2022 | 129 (10.2) | 11.2 | 124 (96.1) | 3 (2.3) | 2 (1.5) | 107 (82.9) | 22 (17.1) | 318 | 2.5 | 69 (53.5) | 7 | 17.8 |
