## Supplementary material for "A Bibliometric Approach to Scientific Production on Familial Hypophosphatemic Rickets in Scopus (2000-2022)": Table 4. Bibliometric indicators of output and impact on journals with fifteen or more articles.

| **Journals (Quartile)** | | **SJR** | **Ndoc** | **NCit** | **Cpd** | **Cited doc (%)** | **H index** | **R index** |
| --- | --- | --- | --- | --- | --- | --- | --- | --- |
| 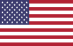 | Journal of Bone and Mineral Research (Q1) | 1.71 | 67 | 4507 | 67.3 | 66 (98.5) | 31 | 63.7 |
| 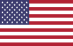 | Bone (Q1) | 1.13 | 55 | 2056 | 37.4 | 55 (100) | 24 | 41.4 |
| 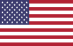 | Journal of Clinical Endocrinology and Metabolism (Q1) | 1.78 | 42 | 2237 | 53.3 | 42 (100) | 22 | 45.4 |
| 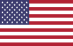 | Calcified Tissue International (Q1) | 0.97 | 34 | 634 | 18.6 | 30 (88.2) | 14 | 22.7 |
| 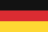 | Journal of Pediatric Endocrinology and Metabolism (Q2) | 0.43 | 31 | 177 | 5.7 | 26 (83.9) | 7 | 10.7 |
| 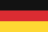 | Pediatric Nephrology (Q1) | 0.77 | 25 | 574 | 23.0 | 22 (88.0) | 13 | 23.1 |
| 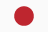 | Clinical Calcium (Non-ranked) | 0 | 24 | 48 | 2.0 | 16 (66.7) | 4 | 5.5 |
| 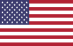 | Endocrinology (Q1) | 1.35 | 18 | 1090 | 60.6 | 18 (100) | 14 | 32.7 |
| 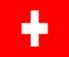 | Frontiers in Endocrinology (Q1) | 1.28 | 17 | 117 | 6.9 | 10 (58.8) | 6 | 10.3 |
| 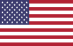 | American Journal of Physiology - Renal Physiology (Q1) | 1.41 | 16 | 862 | 53.9 | 16 (100) | 11 | 28.8 |
| 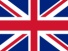 | Orphanet Journal of Rare Diseases (Q1) | 1.12 | 15 | 239 | 15.9 | 12 (80.0) | 7 | 15.1 |
